# Evaluation of Large Language Models in Medical Examinations: A Scoping Review Protocol

**DOI:** 10.1101/2025.06.11.25329442

**Authors:** Weiqi Wang, Baifeng Wang, Yan Zhu, Zhe Wang, Suyuan Peng

## Abstract

**Introduction:** Large language models (LLMs) demonstrate human-level performance in three key domains: linguistic understanding, knowledge-based reasoning, and complex problem-solving. These characteristics make LLMs valuable tools for medical education. Standardized medical examinations evaluate clinical competencies in trainees. These examinations allow rigorous verification of LLMs’ accuracy and reliability in medical contexts. Current methods use standardized examinations to test LLMs’ clinical reasoning abilities. Significant performance variations emerge across different clinical scenarios. No comprehensive reviews have compared different LLM versions in medical examinations. Most studies focus on individual models, lacking comparative analyses of multiple LLM versions. Current approaches struggle to keep pace with evolving research needs. This study synthesizes extant research on LLMs in medical examinations, by analyzing the current challenges and limitations, offers guidance for future investigations.

**Methods and analysis:** The protocol was designed following the *JBI Manual for Evidence Synthesis* guidelines. We established explicit inclusion/exclusion criteria and search strategies. Systematic searches were performed in PubMed and Web of Science Core Collection databases. The methodology details literature screening, data extraction, analysis frameworks, and process mapping. This approach ensures methodological rigor throughout the research process.

**Ethics and dissemination:** This protocol outlines a scoping review methodology. The study involves systematic synthesis and analysis of published literature. It does not include human/animal experimentation or sensitive data collection. Ethical approval is not required for this literature-based study.

**Strengths and limitations of this study:**

1. This scoping review programme strictly adheres to the standardized guidelines for the implementation of scoping reviews. Includes the JBI Manual for Evidence Synthesis and the Preferred Reporting Items for Systematic Reviews and Scoping Reviews Extended Meta-Analysis (PRISMA-ScR) guideline.
2. The search strategy included two databases:PubMed, Web of Science Core Collection.
3. This scoping review will bridge the knowledge gap of LLMs across medical examinations due to recent rapid technological advances.
4. By the nature of the scoping review, failure to critically evaluate identified sources of evidence.

The results of the scoping review will serve as a basis for identifying directions for further research on LLMs in the field of medical examinations.

## Introduction

Recently, the swift advancement of Artificial Intelligence (AI) technologies, notably Large Language Models (LLMs), has ushered in fresh prospects for medical education and examination. These models, trained with deep learning architectures and large-scale corpora, are able to generate, understand, and process natural language texts, and have demonstrated near-human level capabilities in language comprehension and intellectual reasoning. LLMs have emerged as powerful tools across diverse educational domains, particularly in professional fields such as medicine, law, and business^[1]^, where they facilitate both specialized knowledge acquisition and statistical software training^[2]^. Their application in medical education has shown especially promising potential.

Medical examinations play a crucial role in the medical education pathway, can assess the readiness of medical students for clinical practice. In recent years, researchers have increasingly utilized standardised tests, such as medical licensing exams in various languages and countries/regions, to evaluate the performance of LLMs and their ability to apply medical knowledge. The ability of LLMs to provide answers to complex queries ^[1]^ has made their potential as an educational tool in the field of medical examinations increasingly apparent.

For example, OpenAI’s ChatGPT powered by the GPT-3.5 and GPT-4 models^[3]^, has achieved significant results in several examinations, including the Neurosurgery Written Board Examinations^[4]^, the UK Radiology Fellowship Examinations^[5]^, and the Dental Licensing Examinations^[6]^. Both GPT-3.5 and GPT-4 exceeded passing thresholds, with GPT-4 achieving particularly notable results. The GPT-4 model further strengthened in the proportion of facts and questions answered correctly with logically consistent reasoning, demonstrating near-human level performance on a variety of professional and academic benchmarks^[7]^.

Similarly, Google Bard (now Gemini), developed by Google, utilizes the Pathways Language Model (PaLM)2 architecture^[3]^. Gemini has exceeded passing thresholds in multiple standardized examinations, including the Family Medicine In-Training Exam^[8]^, the Ophthalmology Knowledge Assessment^[9]^, and the Japanese national dental hygienist examination^[10]^.

Similarly,Microsoft Copilot (formerly Bing Chat) utilizes the GPT-4 architecture, It has demonstrated proficiency in medical examinations, including the Korean Emergency Medicine Board Examination^[11]^ and the Peruvian National Licensing Medical Examination^[12]^. LLM success extends beyond English-language medical examinations. These models have proven effective across multiple languages, countries, and medical specialties. This is evidenced by performance in the Spanish Medical Residency Entrance Examination (MIR)^[13]^ and the Intercollegiate Membership of the Royal College of Surgeons examination^[14]^.

Studies indicate that although LLMs exhibit strong overall performance, they may fall short of meeting performance benchmarks in specialized domains. In the Japanese Society of Radiology Official Board Exam^[15]^. ChatGPT achieved 40.8% accuracy, GPT-4 reached 65%, and Google Bard scored 38.8%. For the American Board of Anesthesiology (ABA) Examination^[16]^, only GPT-4 attained passing scores. It achieved 78% accuracy in basic components and 80% in advanced sections. GPT-3 scored 58% (basic) and 50% (advanced), while Google Bard achieved 47% and 46% respectively.

LLMs demonstrate exceptional performance in handling fill-in-the-blank, short-answer queries, and expository questions^[17]^. The ability to accurately answer multiple questions based on an article or chart is evident. However, the performance was less effective in handling tasks involving analog data, providing detailed written explanations and answers to complex queries^[17]^. Variations in national healthcare regulations, policies, and languages create training gaps, leading to superior performance in English-language contexts.

While existing studies have preliminarily validated the potential of LLMs in medical examinations, their capacity to comprehensively evaluate multilingual, multidisciplinary, and multi-format examinations remains largely unexplored. In addition, the majority of existing research has primarily focused on the performance of the models. There is relatively limited discussion regarding the analysis of the causes of their errors and potential directions for improvement. Therefore, this study aims to construct a comprehensive framework for measuring medical examinations. It evaluates LLMs performance across clinical scenarios. The research explores LLMs applications in medical education and further promote the innovation and development of AI technology in the medical field. Scoping review through this program, answer the following research questions, see **Table 1**. The purpose of this scoping review program is to address the aforementioned questions, provide an overview of recent developments, and identify major trends. It also aims to guide future research by highlighting the open challenges and limitations of current methods.

**Table 1.**
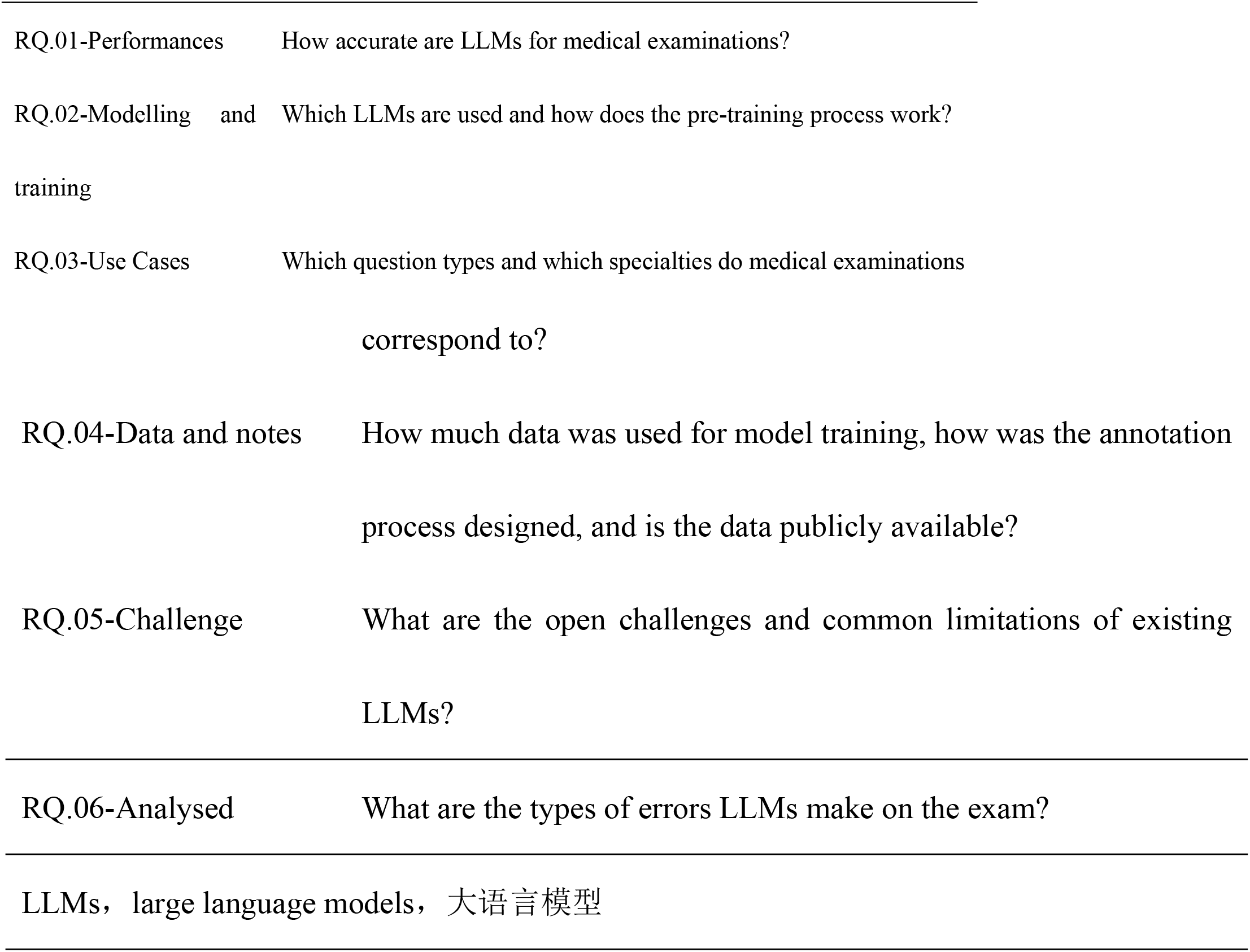
Research sub-questions to be answered based on the scoping review.

## Methods and analysis

A scoping review serves as a systematic synthesis of existing knowledge^[18]^. This methodology aims to address exploratory research questions through comprehensive searching, screening, and evaluation of published literature. It identifies key concepts, evidence categories, and research gaps within specific domains. The scoping review will adhere to the *JBI Manual for Evidence Synthesis*, chapter 11: Scoping reviews. Compliance with the specifications of the Preferred Reporting Items for Systematic Reviews and Meta-Meta Analyses Extended for Scoping Reviews (PRISMA)^[19]^. The study outlines specific methodological components with detailed specifications. It covers the entire process of inclusion and exclusion criteria, literature search strategy, sources of evidence, data extraction, analysis of the evidence, and final presentation of the results. These aspects will be elaborated upon in the subsequent chapters.

### Inclusion exclusion criteria

In **Table 2** and **Table 3**. This section outlines the literature selection criteria.

**Table 2.**
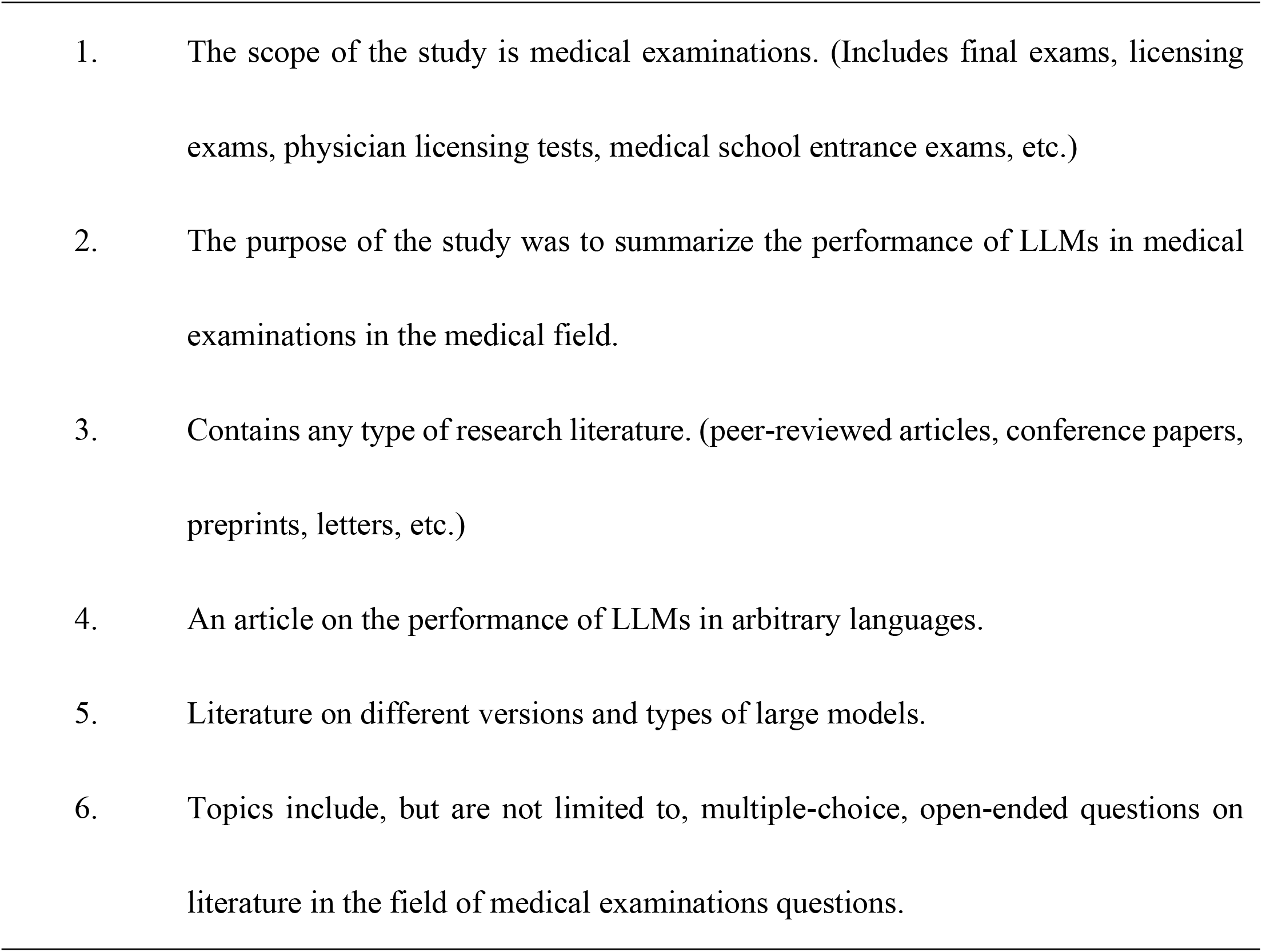
Inclusion criteria.

**Table 3.**
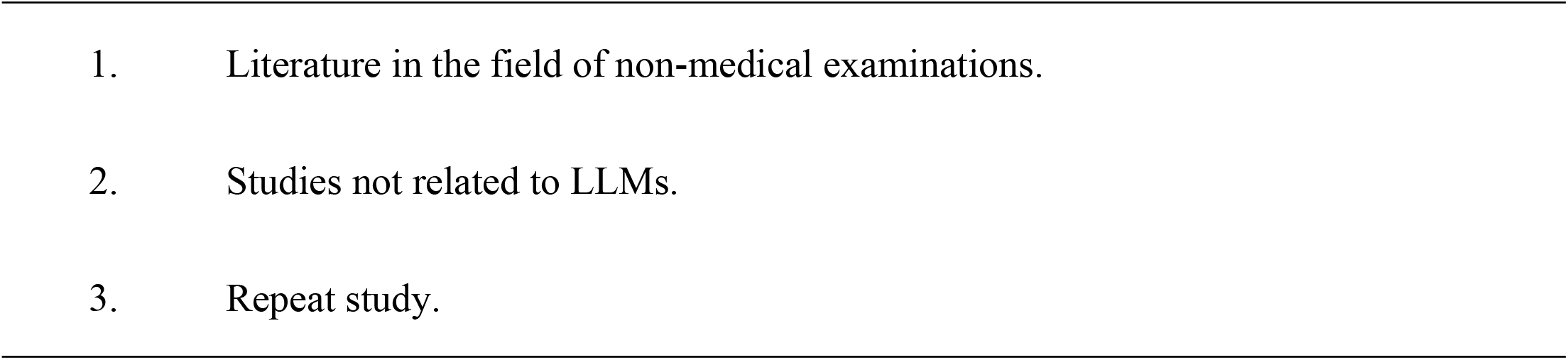
Exclusion criteria.

These criteria are rigorously aligned with the scoping review’s title, primary research questions, and sub-inquiries.

### Search strategy

The search strategy comprised three primary steps. We first conducted preliminary searches in PubMed and Web of Science Core Collection databases. This process identified key terms for search strategy development,as shown in **Table 4**. (The complete search formula used in this study is provided in the Appendix.) Second, we developed systematic search queries through iterative refinement of these terms.

**Table 4.**
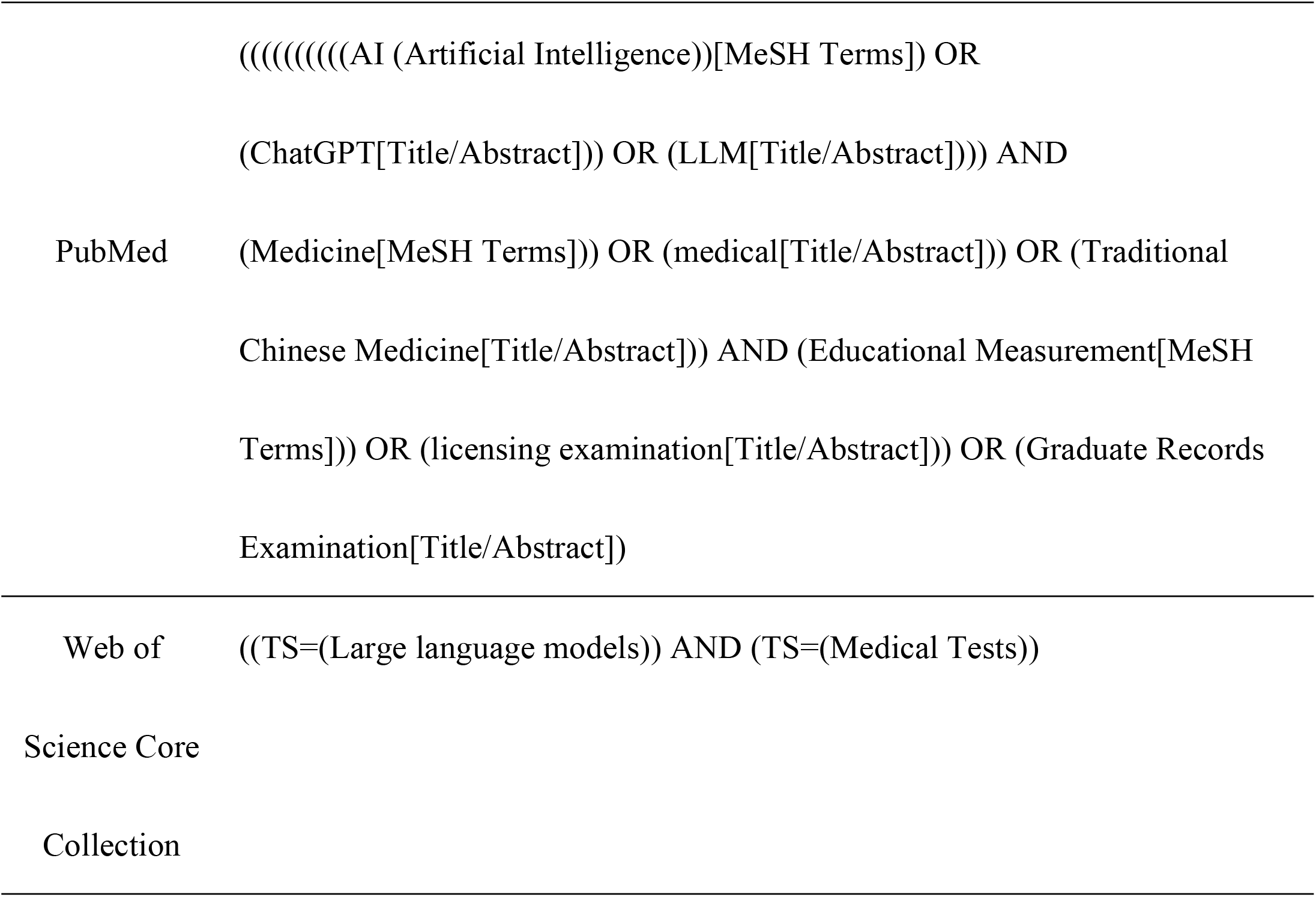
Primary Search Terms.

The search query we employed encompassed two databases: PubMed and the Web of Science Core Collection. Initially, the search queries were primarily focused on PubMed and were subsequently adapted to ensure compatibility with the other database. The aim was to automate the process as much as possible. The literature search for this study was restricted to scholarly publications dated from January 1, 2020, to the present.The delineation of this time window is based on the following two considerations:First, by concentrating on literature published within the past five years, we can effectively capture the cutting-edge developments in the discipline while avoiding the potential timeliness bias that may arise from extending the time span; Second, considering that the medical AI field is characterized by a short cycle of technology updates. The five-year framework strikes a balance between the comprehensiveness of knowledge updates and the feasibility of systematic reviews, ensuring the incorporation of the most recent evidence while maintaining the clarity and operability of research boundaries. The details of each search string - including the number of retrieved records, date range, and search date will be documented using the standardized template established by the Karolinska Institute^[20]^.

Following the study selection process, a forward search (SOE) will be conducted on the reference lists of included articles to ensure comprehensive coverage.This procedure aims to identify potentially relevant studies overlooked during the preliminary search. Systematic examination of references from selected studies will identify theme-related literature and expand the research scope.

### Source of evidence

Evidence source selection defines appropriate literature types and data sources for inclusion. The finalized search strategy will be implemented across both databases. Following search completion, results were deduplicated to eliminate redundant entries. An initial screening phase was conducted on retrieved records. This screening involved title/abstract evaluation against predefined inclusion/exclusion criteria. Studies satisfying all inclusion criteria during initial screening were retained for full-text review. Studies passing initial screening underwent full-text appraisal. Full-text versions were subjected to in-depth evaluation for inclusion criteria compliance. Non-compliant studies were systematically excluded during full-text evaluation. This rigorous screening process yielded a final study cohort. The resultant corpus forms the basis for subsequent analytical procedures. **Figure 1** illustrates the process described.

**Figure 1.**
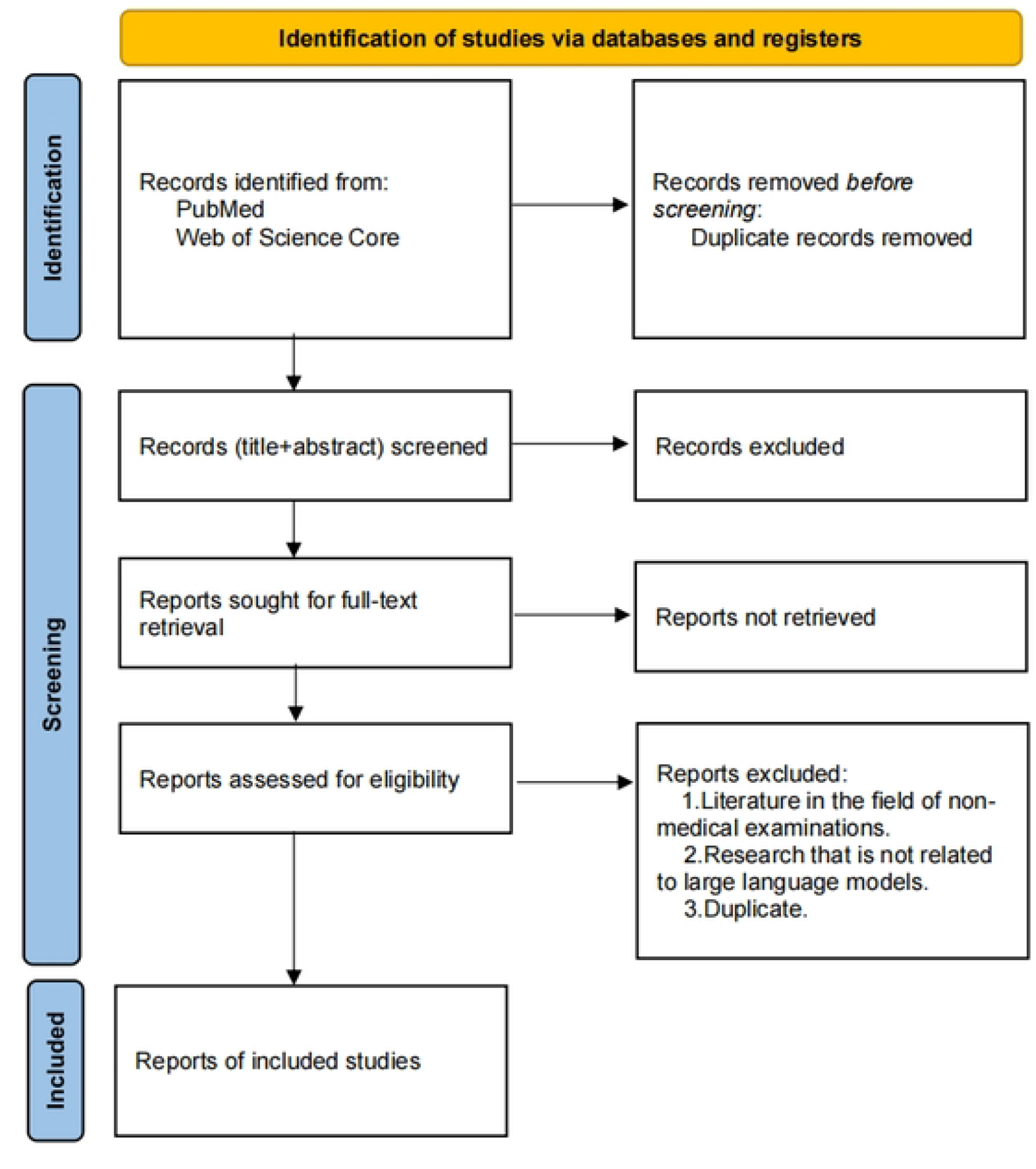
Evidence source selection process.

### Data extraction

Key information was systematically extracted from the included literature. A data charting table was developed following the *JBI Manual for Evidence Synthesis*. The template was modified to align with specific review objectives and subdomains. Details such as: authors, overview, objectives, conclusions, and research methodology. as shown in **Box 1**.

**Box 1.**
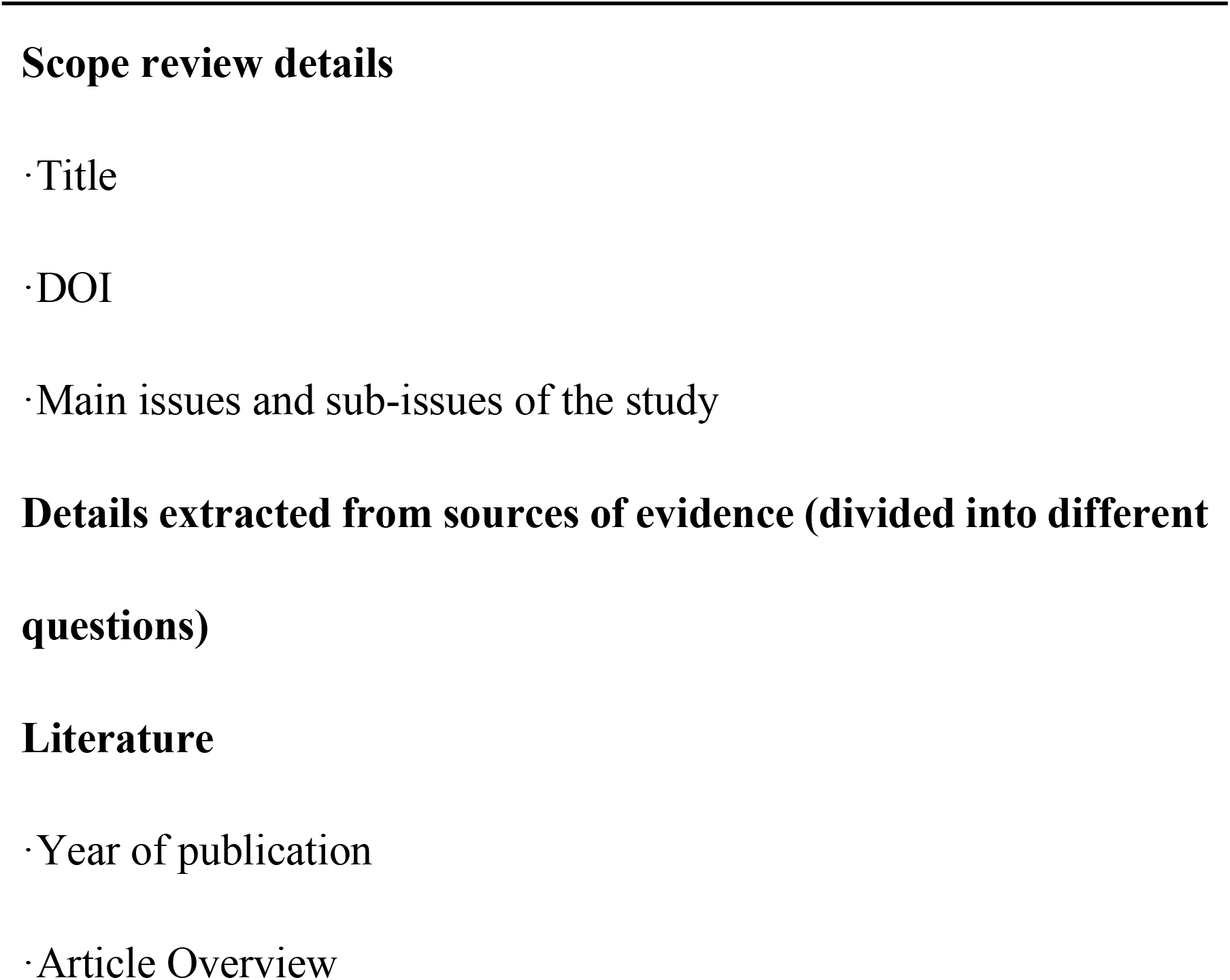
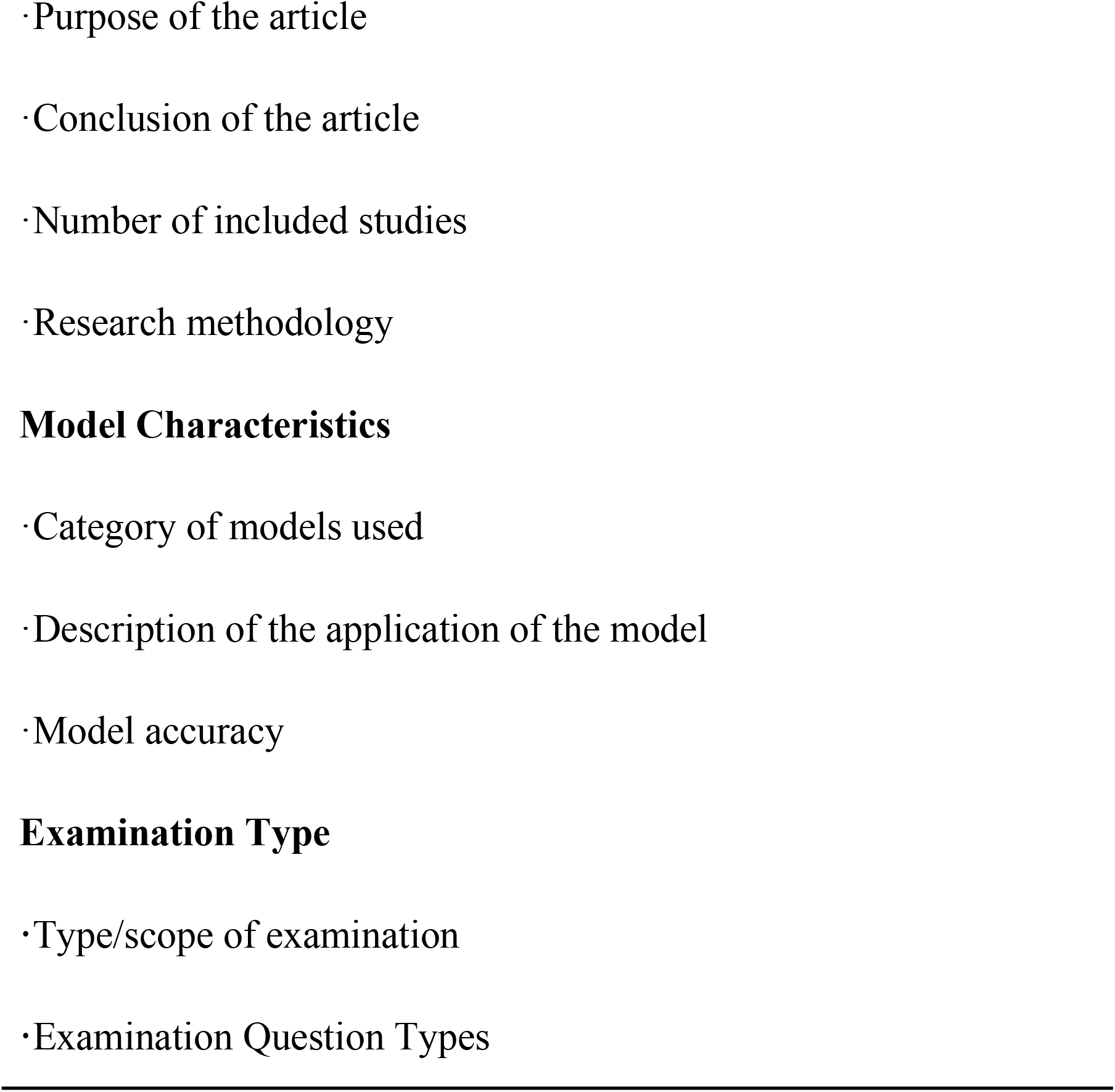
Data charts.

### Analysis of evidence and presentation of results

Descriptive analysis was employed to systematically categorize the compiled evidence. This methodological approach aims to establish a comprehensive overview of current research status. Graphical data visualization techniques were implemented to evaluate LLMs accuracy in medical examination contexts. Percentage as the main quantitative indicator. This presentation not only visualizes the difference in performance between different LLMs across examination. It also provides a data base for exploring the sub-questions in the study (see **Table 1** for details). Annotated graphical elements with explanatory captions ensure precise data interpretation. These annotations provide contextual framework, data significance clarification, and standardized interpretation guidelines.

### Patient and public involvement statement

None.

### Ethics and dissemination

This protocol excludes human/animal subjects research and associated data, thus requiring no ethical approval. Following the announcement of the scoping review agreement, the scoping review itself commences.Final findings will undergo open-access publication in peer-reviewed medical education journals. Any changes to this protocol will be documented, including the date of the change and the reason for the change.

## Data Availability

No datasets were generated or analysed during the current study. All relevant data from this study will be made available upon study completion.

## Patient and public involvement

The design, implementation, reporting, and dissemination plan for this study were conducted without the involvement of patients and/or the public.

## Provenance and peer review

Not applicable.

## Attachment

### Search Formula

#### PubMed

((((((((((((((“Artificial Intelligence”[Mesh]) OR (Artificial Intelligence[Title/Abstract])) OR (Computer Reasoning[Title/Abstract])) OR (AI (Artificial Intelligence[Title/Abstract]))) OR (Machine Intelligence[Title/Abstract])) OR (Computational Intelligence[Title/Abstract])) OR (ChatGPT[Title/Abstract])) OR (LLM[Title/Abstract])) OR (GPT[Title/Abstract])) OR (ERNIE Bot[Title/Abstract])) OR (ChatGLM[Title/Abstract])) AND (((((((((“Medicine”[Mesh]) OR (Medicine[Title/Abstract])) OR (Medical Specialties[Title/Abstract])) OR (Medical Specialty[Title/Abstract])) OR (Medical Speciality[Title/Abstract])) OR (Medical Specialities[Title/Abstract])) OR (nursing[Title/Abstract])) OR (medical[Title/Abstract])) OR (Traditional Chinese Medicine[Title/Abstract]))) AND (((((((((((((((“Educational Measurement”[Mesh]) OR (Educational Measurement[Title/Abstract])) OR (Educational Measurements[Title/Abstract])) OR (Graduate Records Examination[Title/Abstract])) OR (Graduate Records Examinations[Title/Abstract])) OR (Occupational Therapy[Title/Abstract])) OR (licensing examination[Title/Abstract])) OR (medical licensing examination[Title/Abstract])) OR (biomedical science exams[Title/Abstract])) OR (Medical Residency Examination[Title/Abstract])) OR (dental hygienist examination[Title/Abstract])) OR (Nursing Licensure Examinations[Title/Abstract])) OR (standardized examinations[Title/Abstract])) OR (Pharmacy licensure[Title/Abstract])) OR (NMLE[Title/Abstract]))))

#### Web of Science

((((TS=(Large language models)) OR TS=(ChatGPT)) AND TS=(Medical Tests)) AND TS=(Medical Examination))

## Supporting information

**S1 Table. Research sub-questions to be answered based on the scoping review**. This is the S1 Table legend.

**S2 Table. Inclusion criteria**. This is the S2 Table legend.

**S3 Table. Exclusion criteria**. This is the S3 Table legend.

**S4 Table. Primary Search Terms**. This is the S4 Table legend.

**S1 Fig. Evidence source selection process**. This is the S1 Fig legend.

**S1 Box. Data charts**. This is the S1 Box legend.

## REFERENCES

[1] Liu MX, Okuhara T, Chang XY, et al. Performance of ChatGPT Across Different Versions in MedicalLicensing Examinations Worldwide:Systematic Review andMeta-Analysis [J]. Journal of Medical Internet Research, 2024, 26.

[2] Meo AS, Shaikh N, Meo SA. Assessing the accuracy and efficiency of Chat GPT-4 Omni (GPT-4o) in biomedical statistics: Comparative study with traditional tools [J]. Saudi Med J, 2024, 45(12): 1383–1390.

[3] Du W, Jin X, Harris JC, et al. Large language models in pathology: A comparative study of ChatGPT and Bard with pathology trainees on multiple-choice questions [J]. Ann Diagn Pathol, 2024, 73: 152392.

[4] Ali R, Tang OY, Connolly ID, et al. Performance of ChatGPT and GPT-4 on Neurosurgery Written Board Examinations [J]. Neurosurgery, 2023, 93(6): 1353–1365.

[5] Ariyaratne S, Jenko N, Mark Davies A, et al. Could ChatGPT Pass the UK Radiology Fellowship Examinations? [J]. Acad Radiol, 2024, 31(5): 2178–2182.

[6] Chau RCW, Thu KM, Yu OY, et al. Performance of Generative Artificial Intelligence in Dental Licensing Examinations [J]. International Dental Journal, 2024, 74(3): 616–621.

[7] Achiam OJ, Adler S, Agarwal S, et al. GPT-4 Technical Report, F, 2023 [C].

[8] Hanna RE, Smith LR, Mhaskar R, et al. Performance of Language Models on the Family Medicine In-Training Exam [J]. Fam Med, 2024, 56(9): 555–560.

[9] Mihalache A, Grad J, Patil NS, et al. Google Gemini and Bard artificial intelligence chatbot performance in ophthalmology knowledge assessment [J]. Eye (Lond), 2024, 38(13): 2530–2535.

[10] Yamaguchi S, Morishita M, Fukuda H, et al. Evaluating the efficacy of leading large language models in the Japanese national dental hygienist examination: A comparative analysis of ChatGPT, Bard, and Bing Chat [J]. J Dent Sci, 2024, 19(4): 2262–2267.

[11] Lee GU, Hong DY, Kim SY, et al. Comparison of the problem-solving performance of ChatGPT-3.5, ChatGPT-4, Bing Chat, and Bard for the Korean emergency medicine board examination question bank [J]. Medicine (Baltimore), 2024, 103(9): e37325.

[12] Torres-Zegarra BC, Rios-Garcia W, Ñaña-Cordova AM, et al. Performance of ChatGPT, Bard, Claude, and Bing on the Peruvian National Licensing Medical Examination: a cross-sectional study [J]. J Educ Eval Health Prof, 2023, 20: 30.

[13] Guillen-Grima F, Guillen-Aguinaga S, Guillen-Aguinaga L, et al. Evaluating the Efficacy of ChatGPT in Navigating the Spanish Medical Residency Entrance Examination (MIR): Promising Horizons for AI in Clinical Medicine [J]. Clin Pract, 2023, 13(6): 1460–1487.

[14] Chan J, Dong T, Angelini GD. The performance of large language models in intercollegiate Membership of the Royal College of Surgeons examination [J]. Ann R Coll Surg Engl, 2024, 106(8): 700–704.

[15] Toyama Y, Harigai A, Abe M, et al. Performance evaluation of ChatGPT, GPT-4, and Bard on the official board examination of the Japan Radiology Society [J]. Jpn J Radiol, 2024, 42(2): 201–207.

[16] Angel MC, Rinehart JB, Cannesson MP, et al. Clinical Knowledge and Reasoning Abilities of AI Large Language Models in Anesthesiology: A Comparative Study on the American Board of Anesthesiology Examination [J]. Anesth Analg, 2024, 139(2): 349–356.

[17] Stribling D, Xia YX, Amer MK, et al. The model student: GPT-4 performance on graduate biomedical science exams [J]. Scientific Reports, 2024, 14(1).

[18] Hynes H, Wiese A, McCarthy N, et al. International medical graduates’ experiences of clinical competency assessment in postgraduate and licensing examinations: A scoping review protocol [J]. PLoS One, 2024, 19(11): e0305014.

[19] Reichenpfader D, Müller H, Denecke K. Large language model-based information extraction from free-text radiology reports: a scoping review protocol [J]. BMJ Open, 2023, 13(12): e076865.

[20] Karolinska Institutet. University library. Presenting a search strategy. 2022. Available:https://kib.ki.se/en/search-evaluate/searchinginformation/presenting-search-strategy

